# Capturing longitudinal change in cerebellar ataxia: Context-sensitive analysis of real-life walking increases patient relevance and effect size

**DOI:** 10.1101/2024.10.25.24315906

**Authors:** Jens Seemann, Theresa Beyme, Natalie John, Florian Harmuth, Martin Giese, Ludger Schöls, Dagmar Timmann, Matthis Synofzik, Winfried Ilg

## Abstract

**OBJECTIVES:** With disease-modifying drugs for degenerative ataxias on the horizon, ecologically valid measures of motor performance that can detect patient-relevant changes in short, trial-like time frames are highly warranted.

In this 2-year longitudinal study, we aimed to unravel and evaluate measures of ataxic gait which are sensitive to longitudinal changes in patients’ real life by using wearable sensors.

**METHODS:** We assessed longitudinal gait changes of 26 participants with degenerative cerebellar disease (SARA:9.4±4.1) at baseline, 1-year and 2-year follow-up assessment using 3 body-worn inertial sensors in two conditions: (1) laboratory-based walking (LBW); (2) real-life walking (RLW) during everyday living. In the RLW condition, a context-sensitive analysis was performed by selecting comparable walking bouts according to macroscopic gait characteristics, namely bout length and number of turns within a two-minute time interval. Movement analysis focussed on measures of spatio-temporal variability, in particular stride length variability, lateral step deviation, and a compound measure of spatial variability (*SPCmp*).

**RESULTS:** Gait variability measures showed high test-retest reliability in both walking conditions (ICC > 0.82). Cross-sectional analyses revealed high correlations of gait measures with ataxia severity (SARA, effect size ρ≥0.75); and in particular with patients’ subjective balance confidence (ABC score, ρ≥0.71), here achieving higher effect sizes for real-life than lab-based gait measures (e.g. *SPCmp*: RLW ρ=0.81 vs LBW ρ=0.71).

While the clinician-reported outcome SARA showed longitudinal changes only after two years, the gait measure *SPCmp* revealed changes already after one year with high effect size (r_prb_=0.80). In the subgroup with spinocerebellar ataxia type 1, 2 or 3 (SCA1/2/3), the effect size was even higher (r_prb_=0.86). Based on these effect sizes, sample size estimation for the gait measure *SPCmp* showed a required cohort size of n=42 participants (n=38 for SCA_1/2/3_ subgroup) for detecting a 50% reduction of natural progression after one year by a hypothetical intervention, compared to n=254 for the SARA.

**CONCLUSIONS:** Gait variability measures revealed high reliability and sensitivity to longitudinal change in both laboratory-based constrained walking as well as in real-life walking. Due to their ecological validity and larger effect sizes, characteristics of real-life gait recordings are promising motor performance measures as outcomes for future treatment trials.

## Introduction

Gait measures constitute promising candidates for motor performance outcome measures in upcoming therapeutic intervention studies in ataxias^1, 2^, since gait disturbances often present as the first signs of degenerative cerebellar disease (DCD)^3–5^ and represent one of the most patient-relevant disabling features throughout the disease course^6, 7^. It has been shown in laboratory-based assessments by different movement capture technologies that measures of spatio-temporal variability allow to characterize the specificities of ataxic gait ^8–14^ (for reviews, see ^15–18^) with high sensitivity to ataxia severity in cross-sectional and recently also first longitudinal studies ^19, 20^.

With progress in wearable sensor technology enabling gait recordings in patients’ real life, it was hypothesized that those real-life gait measures could be potentially even more sensitive to disease-specific signatures of ataxic gait impairment compared to clinical and lab settings, due to the complexity and challenges of the environments^21–25^, but also due to the larger amount of available walking bouts^26^. In a first cross-sectional study on real-life gait in degenerative cerebellar ataxia, we have shown that ataxic-sensitive gait measures indeed allow not only to capture the gait variability inherent in ataxic gait in real life; but also to demonstrate high sensitivity to small cross-sectional differences in disease severity, with higher effect sizes in real-life walking compared to clinical gait assessment^27^. However, to serve as ecologically valid patient-focused progression and therapy response outcomes, these gait measures have to prove (i) their sensitivity to individual longitudinal change in a sufficiently short time span realistic for intervention trials^1, 28^, as well (ii) their meaningfulness by anchoring with patient-reported, – as required by the FDA^29^.

Noteworthy, to compare patients’ real-life gait behavior at two measurement time points, the influence of context and environment on gait measures have to be considered. These contextual and environmental factors have been shown to have a significant impact on macroscopic gait characteristics, such as average speed, length of walking bouts, and number of turning movements^25, 30–32^. These gait characteristics will differ for indoor (e.g. in a small apartment) vs. outdoor walking, and in turn will influence several gait measures^33, 34^, as it has been shown in healthy participants as well as for different patient populations (Parkinson’s disease, dementia, multiple sclerosis, cerebral palsy)^34–36^. This holds for general performance measures like mean gait speed and even more for variability measures ^35–37^ (e.g. stride length variability, stride duration variability). The analysis of shorter walking bouts for indoor walking - compared to longer outdoor walking - inherently delivers increased variability measures for both healthy participants and patients^36, 38^.

Thus, if one compares measures of a patient’s gait variability at two time points one year apart, increased variability could potentially be caused not by increased balance disturbances, but rather by mere differences in the context and environmental factors of the recorded gait behavior. Thus, to identify disease progression or treatment-induced changes in longitudinal analyses of real-life gait behaviour, one needs to take into account these contextual and environmental factors, and in particular their influence on macroscopic gait characteristics.

In this study, we performed a longitudinal analysis of baseline, one- and two-year follow-up gait recordings in lab-based gait assessment as well as in patients’ individual real life. Matching of longitudinal walking bouts (baseline and follow-up assessments) was performed according to macroscopic characteristics of walking behavior, namely the bout length and number of turns. We hypothesized that gait measures capturing longitudinal change in patients’ real life could be (i) more sensitive to progression in short, trial-like time-frames (e.g. 1 year) compared to lab-based gait assessments and clinical rating scales; and (ii) more patient-relevant in terms of correlation with patient-reported outcomes of balance confidence in important activities of everyday living. This would be key for future treatment trials, as the targeted primary outcome is usually slowing of disease progression in a limited study period, ideally capturable within one year; and by outcomes reflecting patient relevance, as emphasized by the FDA^29^.

## Methods

### Participants and clinical outcome assessments

*Study participants.* 26 participants at an ataxic or preataxic stage of degenerative cerebellar disease (DCD, age: 48±9.5 years) were recruited from the Ataxia Clinics of the University Hospitals Tübingen and Essen. They consisted of 21 participants in the ataxic stage of DCD as defined by a Scale for the Assessment and Rating of Ataxia [SARA] score of ≥ 3 (group ATX; SARA: 9.4±3.2 points), and 5 participants with repeat-expansions in SCA2, SCA3, or SCA6 at the preataxic stage of DCD (SARA score <3) (group PRE; SARA: 1.6±0.65 points)^39^. A total of 18/26 DCD participants carried a repeat expansion in SCA1,2, or 3 (SCA_1/2/3_ subgroup). All main analyses were additionally performed in this subgroup, as these fast-progressing polyQ SCA types are a common promising target in many upcoming intervention trials^1, 2^. Details of patient characteristics are shown in Table 1.

**Table 1.**
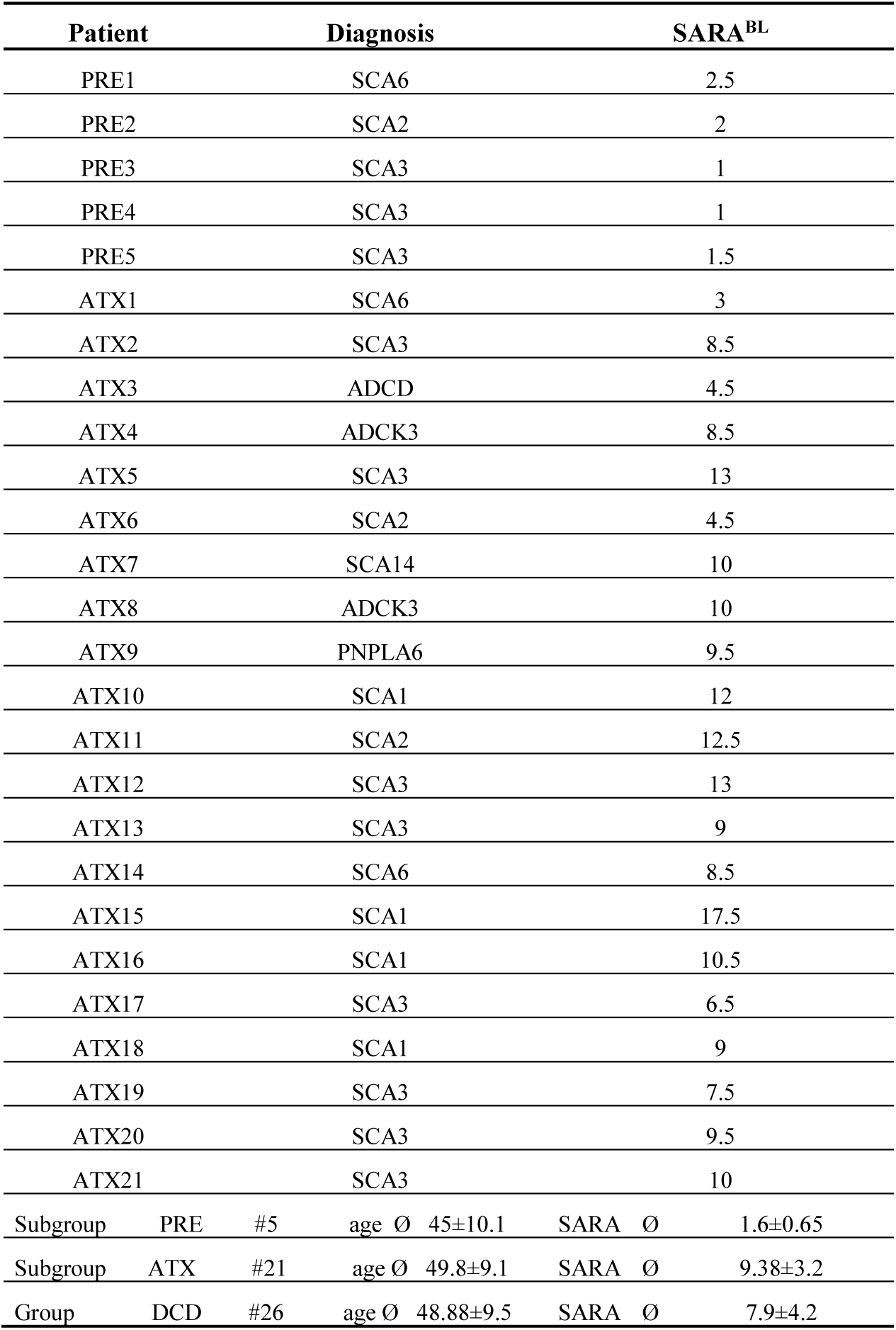
Patient characteristics at baseline assessment. Clinical ataxia severity was determined by the SARA^39^. ADCD: autosomal dominant ataxia of still undefined genetic cause; SCA: autosomal-dominant spinocerebellar ataxia of defined genetic type. The following diagnosis denotes the gene underlying the respective ataxia type: ADCK3 (=ARCA 2, Autosomal-recessive cerebellar ataxia type 2); PNPLA6.

Patients were included based on the following inclusion criteria: 1.) degenerative cerebellar ataxia in the absence of any signs of secondary CNS disease; 2.) age between 18 and 75 years; 3.) able to walk without walking aids. Exclusion criteria were: severe visual or hearing impairment, cognitive impairment (as assessed by the clinician global impression as standard part of the INAS^40^, particularly in relation to limitations in understanding instructions or performing the gait task), or orthopaedic limitations (e.g., severe arthrosis, or previous lower limb fractures or hip/knee replacements) that functionally affect gait. In addition, we recruited 34 healthy controls (HC, age:44.08±14.78 years). Healthy participants had no history of any neurological or psychiatric disease, no family history of neurodegenerative disease, and did not show any neurological signs upon clinical examination. Participants were analyzed cross-sectionally at baseline and, where available, longitudinally at one-year and two-year follow-ups.

#### Clinical outcome assessments and patient-reported outcomes

The severity of ataxia was rated using the SARA^39^. The 3 items rating gait and posture are grouped by the subscore SARA posture & gait (SARA_p&g_)^14, 41^. SARA assessments were performed by expert ataxia neurologists (MS, LS, DT). To capture the impact of disease on subjective confidence in daily living, DCD participants were asked to self-report their balance confidence in activities important in daily living using the Activity-specific Balance Confidence Scale (ABC)^42^ .

### Standard protocol approvals, registration, and patient consent

The experimental procedure was approved by the local ethics committee (598/2011BO1; 303/2008BO2). All participants gave their informed consent prior to participation.

### Gait Conditions

Walking movements were recorded in two different conditions, namely: (i) **L**ab-**B**ased **W**alking (LBW condition): Participants walked 60m straight on a 30m indoor floor (i.e. including one turn) at their preferred self-selected speed on a pre-specified straight route in an institutional setting, supervised without any distractions. The turn is excluded from the analysis. (ii) **R**eal-**L**ife **W**alking (RLW condition): unconstrained walking during participants’ usual individual everyday living, where participants were free to move how they wanted and were used to in their individual daily life, without supervision by any study personnel (total recording time: 4-6 hours). Participants were instructed to wear the sensors inside and outside their home, and include at least a half-hour walk. Participants were instructed to wear the sensors in consecutive recording sessions, each with a duration of maximal 2 hours. Participants documented their recorded walking movements in an activity protocol.

### Movement recordings and gait measures

Three Opal inertial sensors (APDM, Inc., Portland, US) were attached on both feet and posterior trunk at the level of L5 with elastic Velcro bands. Inertial sensor data was collected and wirelessly streamed to a laptop for the automatic generation of gait and balance metrics by Mobility Lab software (APDM, Inc., Portland, US). For the real-life condition (RLW), data was logged on board of each OPAL sensor and downloaded after the session. Step events, as well as spatio-temporal gait measures from the IMU sensors, were extracted using APDM’s mobility lab software (Version 2)^43^, which has been shown to deliver good-to-excellent accuracy and repeatability^44, 45^.

From the rich source of gait measures, we here adopted a hypothesis-driven approach, focusing on those measures that have been considered promising candidate gait measures in previous work. Recent longitudinal studies from our group in laboratory-based gait analysis identified one-year longitudinal gait change in a SCA3 cohort^19^ and an early SCA2 cohort^20^, revealing stride length variability (*StrideL_CV_*), lateral body sway, lateral step deviation (*LatStepDev*) and a compound measure of *spatial step variability (SPCmp,* combining *StrideL_CV_* and *LatStepDev*)^27^ as most sensitive to longitudinal change in ataxia severity.

#### SPcmp was determined in two steps

*step one* determines for each of the two parameters (*StrideL_CV_*) and (*LatStepDev*) separately the relative value of an individual subject in comparison to the value range of all participants at baseline (resulting in values between [0-1]). In step 2, that measure out of these two measures was taken for final analysis where the individual’s result showed a larger abnormality (shown by a value nearer to 1), see also Equation 1 and Figure S3 in Supplement E3.

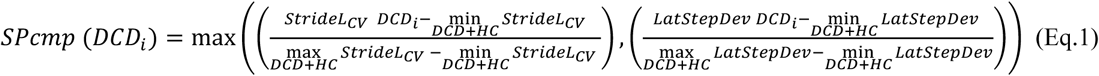

In the real-life walking analysis, we showed that *LatStepDev* and *SPCmp* were sensitive to the cross-sectional ataxia severity^27^, as well as to short-term treatment-related improvements in SCA27B^46^.

In addition, we included gait speed as a general indicator of functional mobility as well as the variability of the lateral angle of the foot during the stance phase, relative to the forward motion of the gait cycle (*ToeOutAngle_SD_*, inspired by^12^). The lateral sway was determined by the coronal range of motion measured by the lumbar sensor (*CorROM_SD_*).

Several variability measures were calculated using the coefficient of variation CV=σ/μ, normalizing the standard deviation with the mean value. On this basis, stride length CV (*StrideL_CV_*) was determined. See detailed description of gait measures in Supplements E2 and E3.

### Selected walking bouts and matching of real-life walking behavior

The analysis focused on walking bouts > 15 strides, as gait variability measures in too short walking bouts are often estimated inaccurately^26^. The first two and last two strides of each bout were removed to reduce the effects of gait initiation and goal-directed deceleration on the variability measures^47^. A walking bout is here defined as a sequence of strides that is not interrupted by a detected turn or a complete halt of at least two median gait cycle durations of the individual. Finally, bouts that showed a jump in gait speed of at least 0.5 m/s between two strides were discarded.

### Matching of real-life walking behavior based on macroscopic gait characteristics

Due to the described influence of contextual and environmental factors on macroscopic gait characteristics - and thus in turn on the examined gait measures-, we introduced a matching procedure for identifying for each subject comparable walking bouts in the longitudinal assessments. This longitudinal matching procedure was performed based on two macroscopic gait descriptors: bout length and number of turns within 60 s before and after the bout (#turns) (see Supplement E1 for details).

To determine test-retest reliability in everyday life, we used the matching procedure described above. Here, the two baseline measurement days with the most bouts were selected and matched. For the lab-based walking task (LBW), we divided the 60m task into two 30m segments (before and after the turn) and calculated the split-half reliability of gait measures (see Statistics). The test-retest reliability of gait measures was calculated using the Intraclass correlation ICC(2, 1)^48^. ICC values <0.5, between 0.5 and 0.75, between 0.75 and 0.9, and >0.90 were considered as poor, moderate, good, and excellent reliability, respectively^48^. Based on the ICC the minimum detectable change (MDC) was calculated, which is critical in determining whether a treatment-related slowing of disease progression can be reliably detected or is lost in the measurement noise^49^.

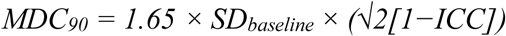

With 1.65 is the z-score of 90 % level of confidence.

### Statistics

Between-group differences (DCD vs. HC group) of movement features were determined by the non-parametric Kruskal-Wallis-test. When the Kruskal-Wallis-test yielded a significant effect (p<0.05), post-hoc analysis was performed using a Mann-Whitney U-test. Effects sizes were determined by Cliff’s delta^50^. Repeated measurement analyses were performed for longitudinal analyses using the non-parametric Friedman test to determine within-group differences between assessments. When the Friedman test yielded an effect (p<0.1), post-hoc analysis was performed using a Wilcoxon-signed-rank-test for pairwise comparisons. Effect sizes r_prb_ for the repeated measurements analyses were determined by matched-pairs rank biserial correlation^51^ and were given with 95% confidence intervals.

We report three significance levels: (i) uncorrected *: p<0.05, (ii) Bonferroni-corrected for multiple comparisons **: p<0.05/n=6: number of analyzed features, (iii) ***p<0.001. Spearman’s ρ was used to examine the correlation between movement measures and SARA scores as well as between measures for different walking conditions. Effect sizes ρ were given with 95% confidence intervals and were classified as ρ: 0.1 small effect, 0.3 medium effect, 0.5 large effect, 0.7 very large effect^52^. Statistical analysis was performed using MATLAB (Version R2024A). Based on the longitudinal changes of the gait measures and the SARA score, a sample size estimation was performed using G*power 3.1^53^ to determine the required cohort size for detecting a 50% reduction of progression by a hypothetical intervention.

### Data Availability

Data will be made available upon reasonable request. The authors confirm that the data supporting the findings of this study are available within the article and its Supplementary material. Raw data regarding human participants (e.g. clinical data) are not shared freely to protect the privacy of the human participants involved in this study; no consent for open sharing has been obtained.

## Results

### Sensitivity of gait measures to ataxia severity: cross-sectional results

Cross-sectional analyses of the baseline assessments revealed group differences between DCD vs HC for several examined gait variability measures in both walking conditions, constrained lab-based walking (LBW) (e.g. *StrideL_CV_*: p=0.002**; *LatStepDev:* p=<0.001***; *SPCmp*: p=<0.001***) as well as real-life walking (RLW) (e.g. *LatStepDev:* p=<0.001***; *SPCmp*: p<0.001***, for an overview of all results see Table 2, Figure 1A). High sensitivity of gait measures *StrideL_CV_*, *LatStepDev,* and *SPCmp* to cross-sectional ataxia severity was indicated by significant correlations with the SARA total score and the SARA_p&g_ subscore, with large effect in both conditions ρ > 0.75, see Table 2. In addition, gait measures revealed high correlations with the patient-reported balance confidence in important activities of daily living, assessed by the ABC score (*StrideL_CV_*, *LatStepDev, SPCmp:* p<0.001***, Table 2). Correlations with the ABC score reveal higher effect sizes for the gait measures in real-life walking (RLW) than lab-based walking (LBW) (e.g. *SPCmp*: ρ=-0.81 in RLW vs ρ=-0.71 in LBW).

**Figure 1.**
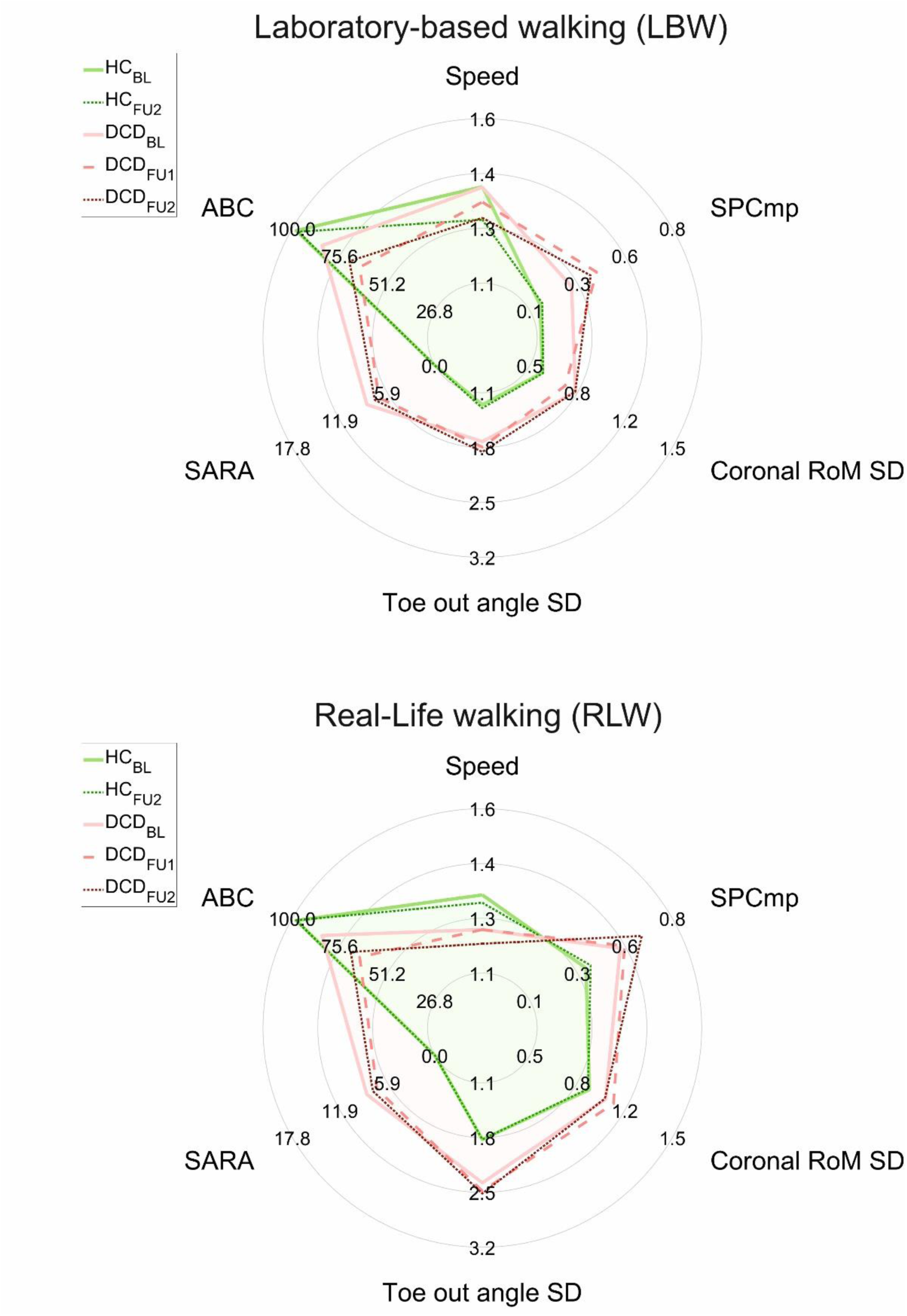
Radar plots illustrating cross-sectional and longitudinal differences on SARA, ABC, and four gait parameters for the gait conditions LBW (A) and RLW (B): Speed, Spatial variability compound measure (*SPCmp*), coronal RoM variability (*CorRoM_SD_*), and foot angle variability (*ToeOutAngle_SD_*). Cross-sectional differences can be seen by comparison of healthy controls (HC_BL_) and the DCD group (DCD_BL_). Given are average values for each group. Longitudinal progression can be seen comparing DCD participants at baseline (DCD_BL_) and 1-follow-up assessments (DCD_FU1_, DCD_FU2_) as well as for healthy controls (HC_BL_ and HC_FU2_).

**Table 2.** Cross-sectional analyses: Between-group differences of healthy controls (HC) and DCD participants for clinician reported outcomes, patient-reported outcomes, and gait measures in the lab-based (LBW) and real-life (RLW) walking conditions. Stars indicate significant differences between groups (*≡ p<0.05, **≡ p<0.0083 Bonferroni-corrected, ***≡ p<0.001). δ denotes the effect sizes determined by Cliff’s delta. Correlations between gait measures and clinician-reported ataxia severity (SARA, SARA_p&g,_) and patient-reported outcomes (ABC) are given for the DCD group. Effect sizes of correlations are given using Spearman’s ρ. CIρ denotes the 95% confidence intervals for ρ. m: mean; sd: standard deviation. Test-retest reliability is analyzed by determining the intraclass correlation coefficient (ICC) (see Methods).

|  |  | group difference<br>DCD vs. HC |  | correlations DCD |  |  |  | ICC |  |
| --- | --- | --- | --- | --- | --- | --- | --- | --- | --- |
| Measure | | p | $\delta$ | SARA | | ABC | | | |
| | | | | $\rho$/CIp | p | $\rho$/CIp | p | | |
| Clinician-reported outcomes | SARA | <0.001*** | 0.99 | - | - | 0.76 | 0.001** | - |  |
|  | SARA<sub>p&g</sub> | <0.001*** | 0.75 | - | - | 0.73 | 0.002** | - |  |
| Patient-reported outcome | ABC | <0.001*** | 0.70 | - | - | - | - | - |  |
| Gait –<br>LBW | Speed | 0.67 | 0.06 | 0.03 [-0.4;0.42] | 0.88 | -0.22 [-0.87;0.34] | 0.45 | 0.94 |  |
|  | StrideL<sub>CV</sub> | 0.002** | 0.47 | 0.79 [0.59;0.90] | <0.001*** | -0.55 [-0.77;0.59] | 0.04* | 0.73 |  |
|  | LatStepDev | <0.001*** | 0.64 | 0.81 [0.54;0.93] | <0.001*** | -0.71 [-0.94;-0.03] | 0.005** | 0.91 |  |
|  | SPCmp | <0.001*** | 0.60 | 0.81 [0.53;0.94] | <0.001*** | -0.71 [-0.91;0.09] | 0.005** | 0.83 |  |
|  | CorRoM<sub>SD</sub> | <0.001** | 0.54 | 0.74 [0.29;0.83] | <0.001*** | -0.72 [-0.85;0.3] | 0.004** | 0.75 |  |
|  | ToeOutAng<sub>SD</sub> | <0.001** | 0.62 | 0.64 [0.41;0.87] | <0.001*** | -0.70 [-0.95;-0.48] | 0.006* | 0.49 |  |
| Gait –<br>RLW | Speed | 0.19 | 0.2 | -0.47 [-0.79;-0.2] | 0.02* | 0.05 [-0.72;0.6] | 0.86 | 0.83 |  |
|  | StrideL<sub>CV</sub> | 0.06 | 0.29 | 0.75 [0.67;0.94] | <0.001*** | -0.35 [-0.91;0.37] | 0.23 | 0.84 |  |
|  | LatStepDev | <0.001*** | 0.61 | 0.77 [0.52;0.93] | <0.001*** | -0.81 [-0.93;0] | <0.001*** | 0.82 |  |
|  | SPCmp | <0.001*** | 0.54 | 0.78 [0.51;0.93] | <0.001*** | -0.81 [-0.92;0.03] | <0.001*** | 0.85 |  |
|  | CorRoM<sub>SD</sub> | 0.13 | 0.23 | 0.59 [0.28;0.81] | 0.002** | -0.62 [-0.98;-0.17] | 0.02* | 0.84 |  |
|  | ToeOutAng<sub>SD</sub> | 0.008** | 0.41 | 0.59 [0.06;0.78] | 0.002** | -0.27 [-0.94;0.33] | 0.36 | 0.79 |  |

Most measures of spatial and temporal variability were highly correlated across the conditions LBW and RLW in cerebellar patients (p<0.003, Supplement E8).

### Gait measures for matched walking bouts show good-to-excellent test-retest-reliability

To identify suitable macroscopic gait characteristics for the matching of walking bouts, linear regression analysis showed significant contributions of both, *bout length* and *number of turns* in explaining the variability for walking bouts of healthy controls (see Supplement E3). Performing the presented matching procedure on the real-life baseline assessment (see Methods), revealed good to excellent test-retest-reliability (ICC) for several gait measures like *LatStepDev*, *StrideL_CV,_* and *SPCmp* (ICC(2,1) ≥ 0.82 (see Table 2).

### Sensitivity of gait measures to longitudinal change

We next analysed whether the gait measures allow to detect longitudinal changes in real life at one-year follow-up (duration: 380±52 days) and at a two-year follow-up assessment (duration: 816±106 days). Longitudinal gait data were available from 23 DCD participants for the first (FU1) and 22 DCD for the second follow-up assessment (FU2). In addition, 34 healthy controls were available for FU1 and 17 for FU2.

Longitudinal analyses revealed for the clinical ataxia score SARA significant changes only in the second follow-up assessment (see Table 3). In contrast, several gait measures - in particular *StrideL_CV_* and the compound measure of spatial variability (*SPCmp*) - revealed significant changes already after one year in the real-life walking condition RLW (*StrideL_CV_*: FU1: p=0.0047**, r_prb_=0.67; *SPCmp*: FU1: p=0.0007***, r_prb_=0.80). The robustness of these results is supported by stable effect sizes in the second follow-up assessment (*SPCmp*: FU2: p=0.016*, r_prb_=0.77) (Figure 2A, Table 3).

**Figure 2.**
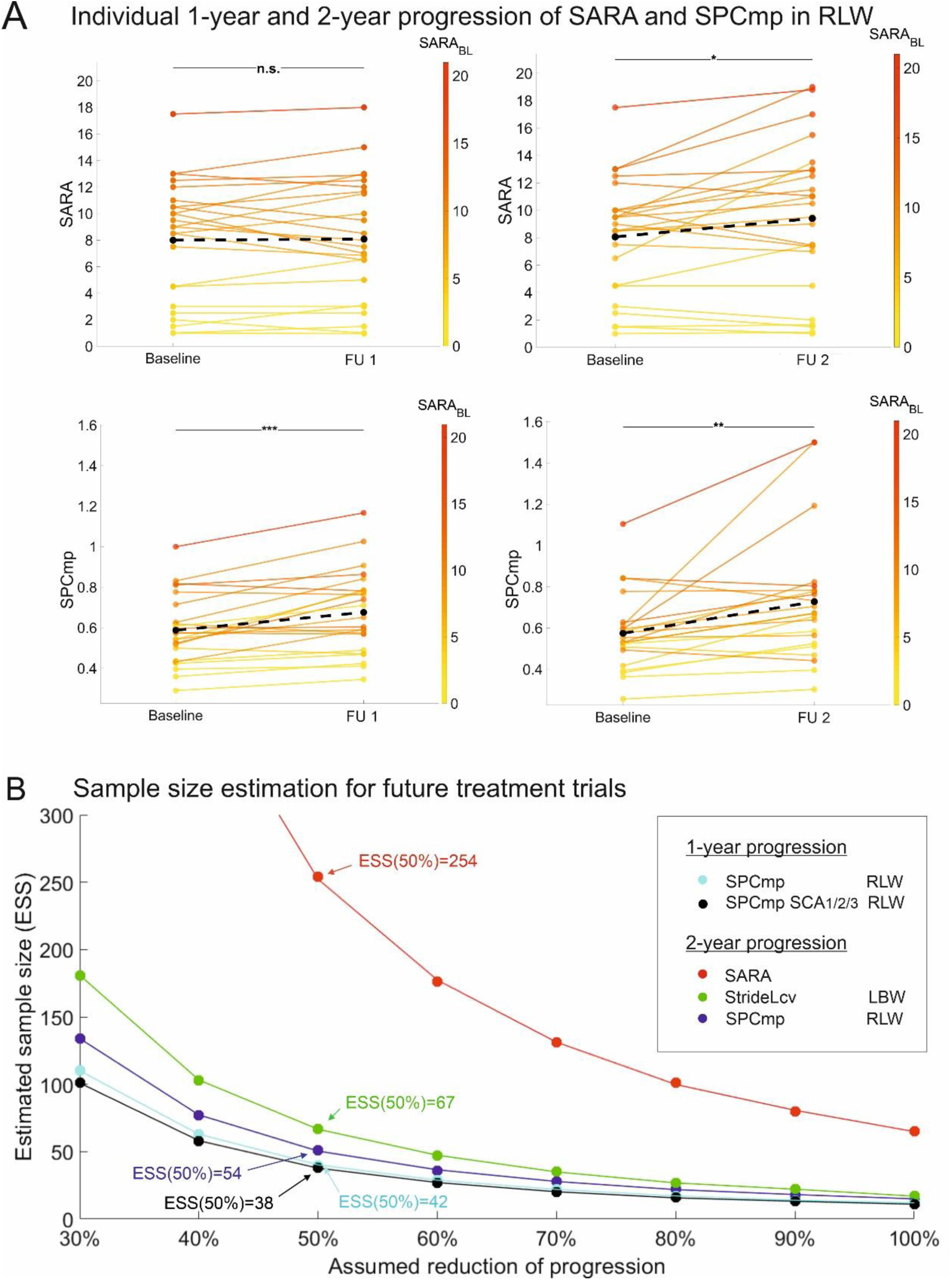
(A) Within-subject changes between baseline and follow-up assessments for the group of DCD participants. Shown are longitudinal differences in SARA (upper panel) and the compound measure SPCmp (lower panels). For both measures, the left panel shows the change between baseline and 1-year follow-up (FU1), and the right panel the change between baseline and 2-year follow-up (FU2). In all panels, SARA scores of individual cerebellar participants are color-coded. Black dotted line = mean change across all participants. Stars indicate significant differences between time points (*≡ p<0.05, **≡ p<0.0083 Bonferroni-corrected, ***≡ p<0.001). (B) Sample size estimates were made for future treatment trials that showed different levels of progression reduction for the various outcome measures: SARA and the gait measures SPCmp and StrideLcv in the laboratory (LBW) and real life (RLW). The estimated number of participants per study arm is plotted against the hypothesized therapeutic effect on reducing 1-year progression or 2-year progression in DCD patients or the SCA_1/2/3_ subgroup (SCA_1/2/3_), respectively.

**Table 3.**
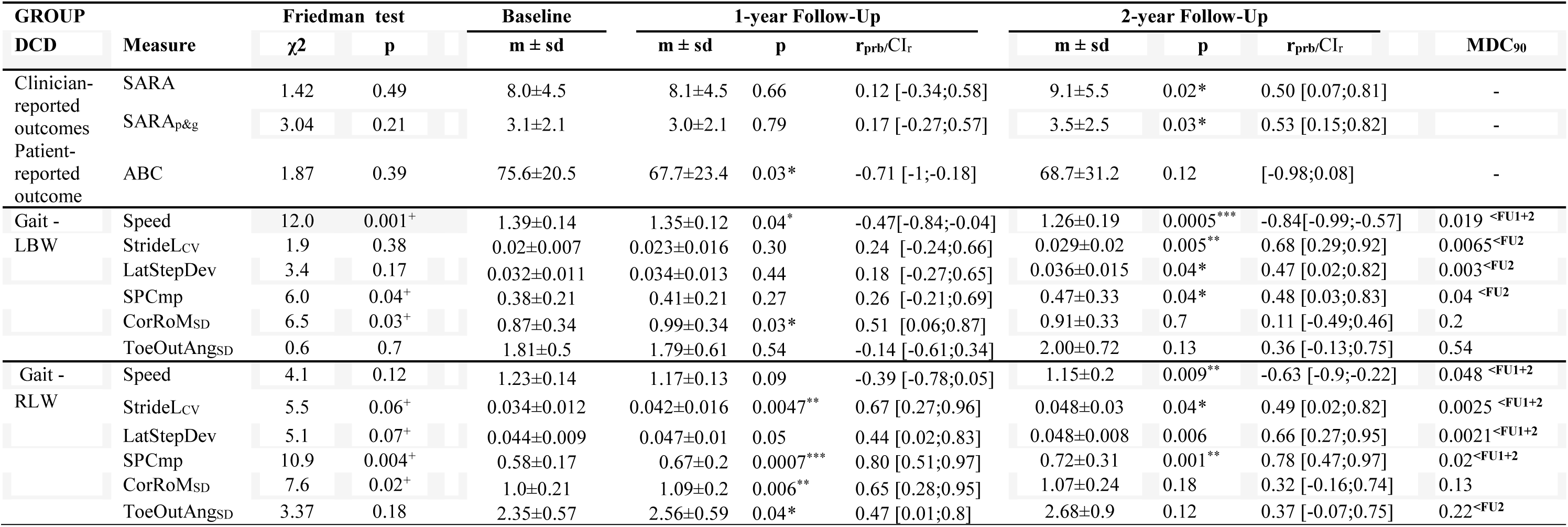
Longitudinal within-subject comparison for the group of degenerative cerebellar disease (DCD). Shown are results of clinician-reported ataxia ratings (SARA score and SARA_p&g_ posture&gait subscore^41^) as well as of gait measures in clinical assessment (LBW) and in real-life walking (RLW) for baseline, 1-year, and 2-year follow-up assessments. Friedman test determined within-group longitudinal differences (^+^, p<0.1). Post-hoc test **p-**values determined by Wilcoxon signed-rank test for both follow-up assessments relative to baseline. Stars indicate significant differences between groups (*≡ p<0.05, **≡ p<0.0083 Bonferroni-corrected, ***≡ p<0.001). Effect sizes r_prb_ determined by matched-pairs rank-biserial correlation^51^. CI_r_ denotes the 95% confidence intervals for effect sizes r_prb_. Shown are analyses for the whole group of degenerative cerebellar disease (DCD), both consisting of preataxic and ataxic participants. MCD_90_ denotes the smallest reliable detectable change (90% confidence interval). The MDC column indicates whether this is smaller than the change between baseline and 1-year Follow-up (‘^<FU1’^) or/and between baseline and 2-year Follow-up (^<FU1+2^).

Moreover, these longitudinal changes already after one year for the gait measures *StrideL_CV_* and *SPCmp* can also be observed and validated in the subgroup SCA_1/2/3_ (FU1: n=18; FU2:n=18) (*SPCmp*: FU1: p=0.0023**, r_prb_=0.86; FU2: p=0.0067**, r_prb_=0.76) – in fact with higher effect size, despite smaller sample size of participants (Supplement E6). For both the patient group DCD as well as for the subgroup SCA_1/2/3_, these annual changes were larger than the minimum detectable change (MDC, see Tables 3 and supplement E6, e.g. *SPCmp*: Δ_BL vs. FU1_= 0.12 > MDC_90_= 0.02). In the HC group, longitudinal changes were observed in gait speed, but not in the ataxic-specific measures of spatio-temporal variability (see Figure 1).

Based on observed effect sizes in the DCD population, sample size estimation for the compound spatial variability measure (*SPCmp*) showed a required cohort size of n=42 for detecting a 50% reduction of natural progression after one year by a hypothetical intervention (90% power and one-sided 5% type I error) in comparison to n=254 for the SARA score (Figure 2 B). Required cohort sizes were even smaller for the SCA_1/2/3_ subgroup (n=38), given the larger effect size (Figure 2B).

Compared to the matched walking trials, analysis of the non-matched walking trials (bout length >15) also showed significant longitudinal differences, yet with smaller effect sizes (e.g. *SPCmp*: FU1: p=0.0089*, r_prb_=0.62; FU2: p=0.01*, r_prb_=0.58, see Supplement E5). Moreover, the analysis of the non-matched walking bouts revealed a correlation between longitudinal differences in variability measures like *StrideL_CV_* and longitudinal differences in mean bout length (r=0.59, p=0.0034**) (Supplement E1), potentially influencing the observed changes. Such correlations were not observed in the matched analysis (Supplement E1).

## Discussion

This study aimed to test the hypothesis that the longitudinal progression of ataxic-related gait impairments can be reliably and sensitively captured in real-life walking by analysis of walking bouts which are matched according to macroscopic gait characteristics reflecting environmental factors. We showed that real-life gait measures can capture longitudinal change within short trial-like time frames like one year with high effect size, thus outperforming in their sensitivity both clinician-reported outcomes like the SARA as well as lab-based gait measures which show longitudinal change only after two years. These results thus indicate an increased sensitivity as well as ecological validity for real-life gait measures as promising motor performance outcomes in upcoming treatment trials.

### Increased spatio-temporal gait variability as a consistent feature of ataxic gait in laboratory-based assessments

Our findings in the constrained walking condition LBW confirm the results of previous studies from our and other groups with different movement capture technologies^8, 9, 12, 13, 19^. These studies showed that spatio-temporal variability measures like stride length variability, stride time variability, and lateral step deviation in constrained walking conditions serve as reliable and valid measures for cerebellar ataxia and - as demonstrated here for wearable sensors - correlate with gait and posture ataxia severity as well as with patients’ subjective balance confidence in important activities of daily living (ABC-score).

### Measures of ataxic gait in real life: cross-sectional sensitivity to ataxia severity

Although real-life gait is inherently far more variable in both healthy controls and cerebellar patients^38^, several of our gait variability measures (e.g. *StrideL_CV_*, *LatStepDev,* and *SPCmp*), allow to capture the specific gait variability inherent in ataxic gait in real life^27^. The compound measure *SPCmp* -integrating variability in the anterior-posterior as well as in the mediolateral dimension - hereby seems to benefit from capturing different compensation strategies used individually and in diverse stages of degenerative cerebellar disease, and might allow capturing gait ataxia in particular in more advanced disease stages (see Supplement E3)^27^.

Consistent with and validating our previous study^27^ cross-sectional sensitivity of real-life gait measures (e.g. *StrideL_CV_*, *LatStepDev,* and *SPCmp*) for ataxia severity was shown by the high correlation with clinical severity of ataxia (p<0.002**, see Table 2). Even more meaningful for patient relevance and ecological validity, however, is the high correlation between gait measures and patients’ subjective balance confidence in important activities of daily living, as assessed by the ABC score (see Table 2). The high effect sizes of our selected gait measures in their correlation with the ABC score – confirmed and numerically even higher for real-life than lab-based gait assessments (*SPCmp*: RLW ρ=0.81, LBW ρ=0.71) - emphasize the relevance of ataxia-related gait impairments, as captured by body-worn sensors, to patients’ everyday life – which is key to FDA-conform patient-focussed outcome and drug development^29^.

### Matching walking bouts according to bout length and number of turns

The influence of contextual and environmental factors on gait measurements during real-life walking is currently under intense investigation in various movement disorders ^25, 32, 35, 37^. So far, matching or selection procedures based on contextual factors or macroscopic gait characteristics have been used exclusively in cross-sectional studies, e.g. to compare a group of patients with healthy controls in activity monitoring and real-life walking, to differentiate patient subgroups^25^, or to compare patients’ real-life walking behaviour with clinical gait assessments^37, 54^. Moreover, in most approaches, the selection of walking sessions was based on bout length alone.

In contrast, the focus of our study was to investigate a longitudinal matching approach as a novel strategy for longitudinal change analysis, allowing to identify comparable walking bouts at baseline and follow-up visits. As turning movements are an important component of real-life walking behavior, and also typically differ between indoor and outdoor walking^30^, we included the number of turning movements as well as the bout length in our macroscopic characterization of walking behavior to identify comparable walking bouts. The number of turning movements indeed contributes to the macroscopic walking characteristics in explaining step variability, as observed by linear regression analysis of the baseline recordings in healthy controls (Supplement E4). For the longitudinal analyses, the matching procedure ensures that observed effects are not predominantly caused by differences in bout length (Supplement E4). Using the proposed matching procedure, we showed good-to-excellent test-retest-reliability within the baseline assessment, including real-life recordings on different days.

Although the non-matched data showed also significant longitudinal changes (Supplement E7), the results from the matched walking bouts revealed larger effect sizes and were more reliable, given that the matching procedure allowed to substantially reduce the influence of longitudinal changes in bout length as a main confounder (Supplement E1).

Thus, in sum, this approach allows comparison of real-life gait assessments without requiring participants to perform similar gait behaviour in all home assessments or capturing home data over a long period of time (where contextual differences can be assumed to average out). At the same time, it highlights the need also for other future longitudinal gait studies to control for changes in macroscopic walking characteristics like bout length and the number of turnings, as potential confounders for longitudinal analyses.

### Gait measures in real-life walking capture longitudinal change with increased effect size

Using these matching procedures, and consistent with the cross-sectional results, measures of spatio-temporal variability in real-life walking (and in particular the compound measure *SPCmp*) show high responsiveness to change at one year. Their effect size in longitudinal sensitivity to change outperforms the clinician-reported outcome (SARA) and laboratory gait measures.

Importantly, the high test-retest reliability, even in real-life conditions, resulted in an average longitudinal change that was higher than the minimum detectable change (MDC) (see Table 3), which is a critical requirement for reliable detection in clinical trials^55^.

The subgroup SCA_1/2/3_ showed even higher effect sizes and consequently smaller sample size estimates. This finding is consistent with previous studies showing faster progression in SCA 1,2 3^56^ compared to a cross-genotype DCD population, which also includes slower progressing DCD types, e.g. SCA6^57^ and non-PolyQ SCAs^58^.

The reduction in sample size inferred by these digital-motor performance outcomes could be decisive for the feasibility of a treatment trial: whereas trials with e.g. 254 SCA participants per trial arm (as required for SARA as outcome) are almost impossible, 38 SCA participants (as required for the gait performance measure *SPCmp* in SCA_1/2/3_)(Figure2B) are well feasible.

### Limitations

Overall, the present study aimed to explore and longitudinally validate digital-motor sensor measures in real-life in ataxia with a cross-genotype cohort, as this approach allows validation digital-motor measures *across* DCDs (whereas validation within each single genotype would take many additional years and effort). This approach was based on the assumption that our digital-motor measures would capture functional impairment generically across DCDs, given that they qualitatively affect the same ataxia-related functions. This assumption is being corroborated by our previous work demonstrating validity of ataxia-specific gait measures (e.g. *StrideL_CV_*, *LatStepDev,* and *SPCmp*) across various DCDs ^20, 46, 59^; as well as by the comparable longitudinal results for the overall DCD cohort and SCA1/2/3 subgroup observed in the current study. However, given that different DCD genotypes gradually differ in their specific progression rates, the validity of the sensitivity of digital motor measures to detect longitudinal changes formally remains to be demonstrated within specific genotypes as well as pre-ataxic populations only. Larger multi-centre studies focussing on real-life behaviour are thus warranted to confirm our results in larger cohorts sufficiently powered for genotype-specific analyses and including a higher number of preataxic participants. In addition, further work could ideally include longer real-life assessments (e.g. two weeks) to quantify day-to-day variability in patients’ motor behaviour.

## Conclusion

This study unravels methods and measures that allow to quantify longitudinal changes in real-life ataxic gait with high effect size and high correlations with patient-reported outcomes of daily living, thus yielding promising ecologically valid, patient-focussed outcome measure candidates for future natural history and treatment trials in degenerative cerebellar ataxias. In addition to the higher effect sizes gained from real-life assessments, these measures allow for the objective quantification of patients’ real-life motor performance - instead of clinical outcome assessment in partly “artificial” settings and tasks, e.g. by motor tasks as part of clinical scores (=SARA) or under lab conditions (=standard digital-motor assessments), which serve as surrogate parameters at best. Thus, measures of real-life motor performance add ecological validity and thus help to inform upcoming treatment trials in degenerative cerebellar ataxias and FDA-compatible development of patient-focused outcomes and approval of novel treatments^21, 28^.

## Acknowledgments

This work was supported by the International Max Planck Research School for Intelligent Systems (IMPRS-IS) (to J.S.) and the Else Kröner-Fresenius-Stiftung Medical Scientist programme ‘ClinbrAIn’ (to W.I. and M.G.). as well as the Else Kröner-Fresenius Stiftung Clinician Scientist program “PRECISE.net” (to M.S.). In addition, this work was supported by the European Union, project European Rare Disease Research Alliance (ERDERA, # 101156595) (to M.S.).

## Appendix I Authors

| Name | Location | Role | Contribution |
| --- | --- | --- | --- |
| Jens Seemann, MSc | Hertie Institute for Clinical Brain Research, Tübingen, Germany | Author | Execution of the study, analysis and interpretation of the data, revising the manuscript |
| Theresa Beyme | Hertie Institute for Clinical Brain Research, Tübingen, Germany | Author | Execution of the study, analysis of the data, revising the manuscript |
| Natalie John | Hertie Institute for Clinical Brain Research, Tübingen, Germany | Author | Execution of the study, analysis of the data, revising the manuscript |
| Florian Harmuth, MSc | Institute of Medical Genetics and Applied Genomics, University Hospital Tübingen | Author | Execution of the study, analysis of the data, revising the manuscript |
| Martin Giese, PhD | Hertie Institute for Clinical Brain Research, Tübingen, Germany | Author | interpretation of the data, revising the manuscript |
| Ludger Schöls, MD | Hertie Institute for Clinical Brain Research, Tübingen, Germany | Author | interpretation of the data, revising the manuscript |
| Dagmar Timmann, MD | Essen University Hospital University of Duisburg-Essen, Germany | Author | Organisation of the study, interpretation of the data, revising the manuscript |
| Matthis Synofzik, MD | Hertie Institute for Clinical Brain Research, Tübingen, Germany | Author | design and conceptualization of the study, interpretation of the data, drafting the manuscript. |
| Winfried Ilg, PhD | Hertie Institute for Clinical Brain Research, Tübingen, Germany | Author | design and conceptualization of the study, analysis and interpretation of the data, drafting the manuscript. |

## Supplementary information

### Supplement E1- Matching of walking bouts based on bout length and number of turns

For matching, an Euclidean distance matrix D was created for each subject using bout length and #turns for each baseline bout to each follow-up bout (see Figure S1). To weight both criteria equally, the data were previously standardized per subject (centered and normalized to one standard deviation of the participants’ baseline measurement). Using the distance matrix D for each subject as a cost matrix, a linear assignment problem was solved to obtain a 1:1 matching of similar bouts for baseline and follow-up. A cost for non-assignment of 0.5 was set, to obtain bouts that were as similar as possible, but at the same time to avoid losing any participants for the analysis. Thus, for each subject, only bouts with similar macroscopic gait descriptors are compared. Mismatched bouts are not included in the analysis, reducing purely contextual longitudinal differences.

**Figure S1.**
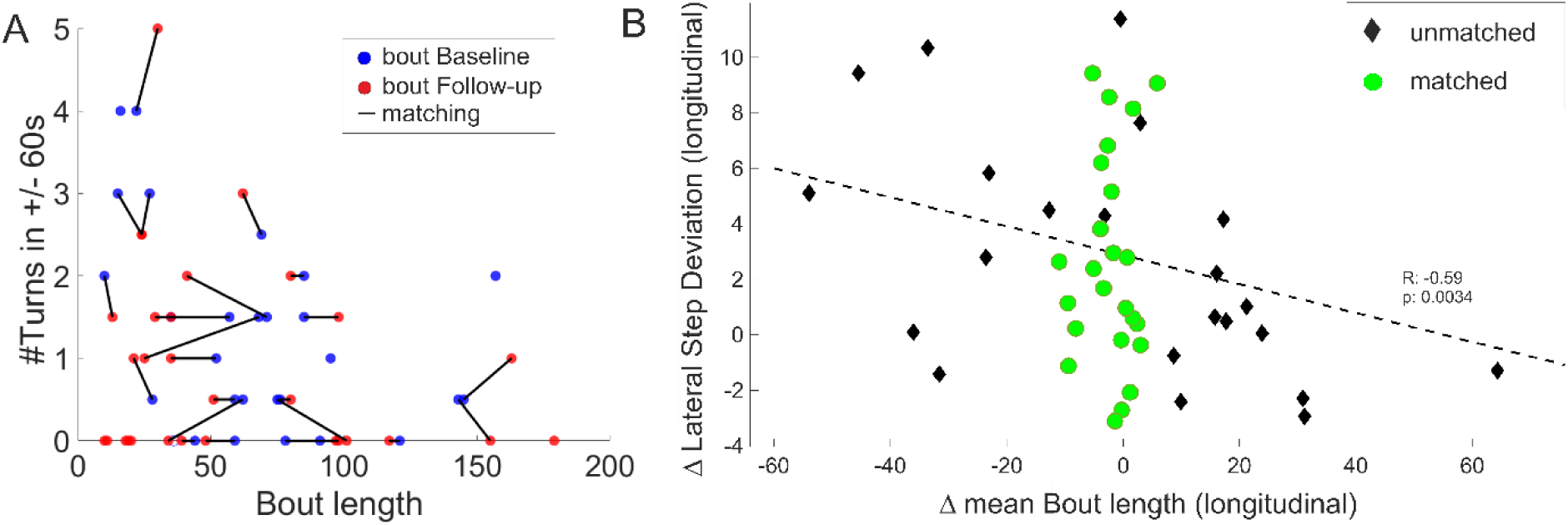
(A) Illustration of the Mapping procedure of corresponding real-life walking trials at baseline (blue) and the one-year follow-up (red). (B) Relationship between longitudinal differences (Δ= one-year follow-up – baseline) differences in mean bout length and Lateral Step deviation for the DCD participants (non-matched: red; matched: blue). In the non-matched condition, there is a significant correlation between the longitudinal difference in Lateral Step Deviation and the difference in mean bout length (r=0.59, p=0.0034**).

### Supplement E2- Details on gait measures

#### Lateral step deviation (*LatStepDev*)

This gait measure was determined based on three consecutive walking steps, calculating the absolute amount of perpendicular deviation of the middle foot placement from the line connecting the first and the third step (Figure S2). *LatStepDev* was normalized with stride length (% of stride length), thus providing a measure independent from stride length variability, which is suggested to be increased in real-life gait.

#### Coronal Range of Motion (*CorRoM*)

The angular range of the thoracic spine in the coronal plane (roll).^12^

#### Toe Out Angle (*ToeOutAng*)

The lateral angle of the foot during the stance phase, relative to the forward motion of the gait cycle. Positive angle is outward rotation.^12^

**Figure S2.**
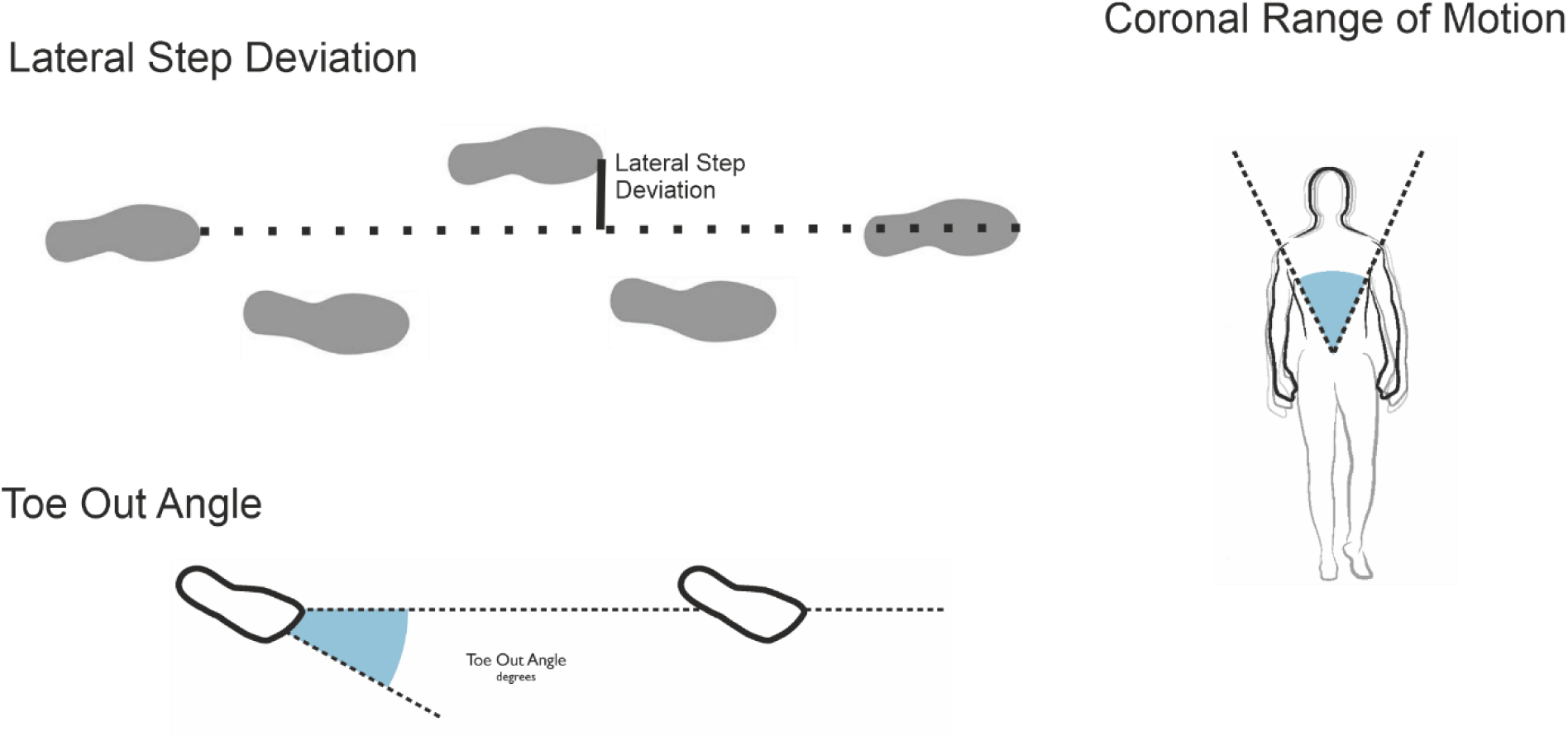
Illustration of the gait measures: Lateral step deviation, Toe Out Angle, and Coronal Range of Motion (Adopted from^12^).

### Supplement E3 – Compound measure of spatial variability

The spatial step variability compound measure *SPCmp was determined in two steps: step one* determines for each of the two parameters (*StrideL_CV_*) and (*LatStepDev*) separately the relative value of an individual subject in comparison to the value range of all participants (resulting in values between [0-1], see Figure S3 A). In step 2, that measure out of these two measures was taken for final analysis where the individual’s result showed a larger abnormality (shown by a value nearer to 1), whereas the respective other measure was not entered into the further analysis, see Figure S3-A. For the longitudinal analysis, the value range of all participants is determined based on the baseline assessment. Therefore, values in the follow-up assessment can exceed the range [0-1].

**Figure S3.**
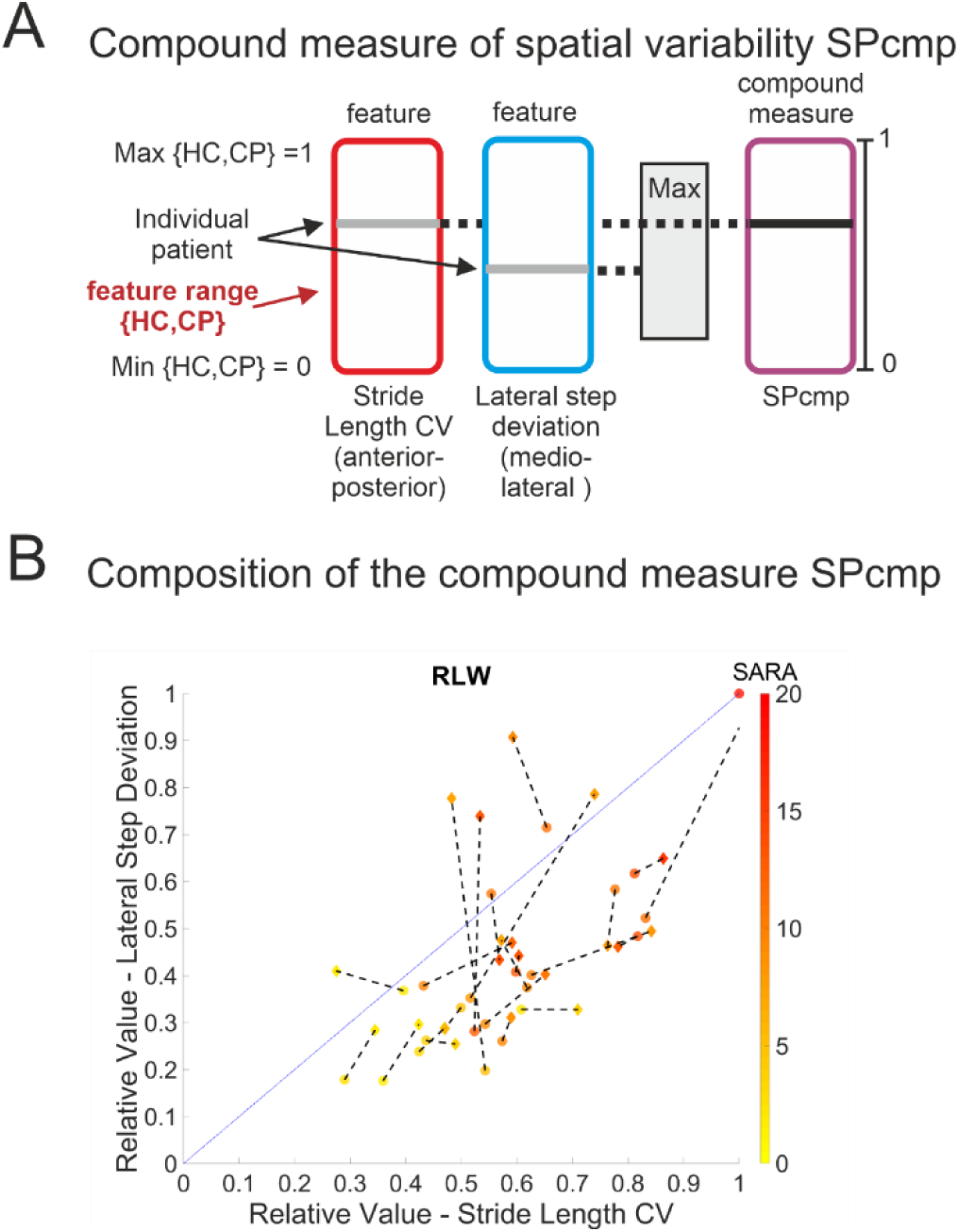
(A) Determination of the compound measure SPCmp. It is determined by the maximum of the relative values for the parameters StrideL_CV_ and LatStepDev. (B) Composition of the compound measure SPCmp for the walking conditions RLW in Baseline and the first follow-up assessment. Shown are the relative parameter values of each patient for the parameters StrideL_CV_ (x-axis) and LatStepDev (y-axis). The color-coding denotes the severity of gait and posture ataxia as determined by the SARA score. Dotted lines connect the baseline and the follow-up examination for the individual patients.

### Supplement E4 Linear regression models of the matching procedure

We performed linear regression models to examine the influence of macroscopic gait characteristics on our variability measures.

#### Linear regression models

The regression models examined the predictability of the gait measures lateral step deviation and stride length from different high-level gait descriptors like bout length, number of turns, SARA, or age of the participants. The regression was conducted on all ataxic and pre-ataxic participants in the baseline and follow-up condition, where each bout represented one data point.

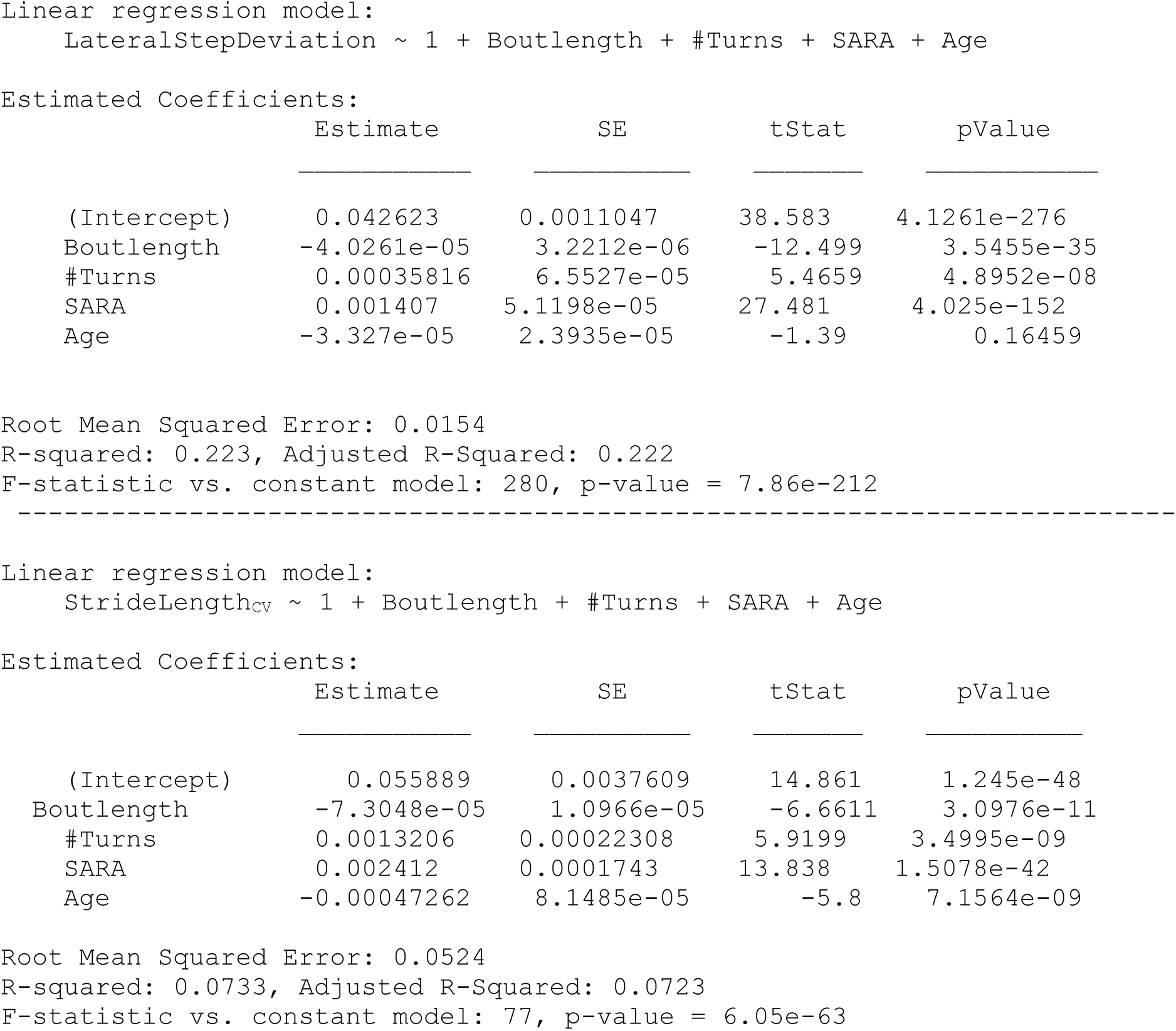

### Supplement E5 – Comparison of statistics between matched and non-matched walking bouts

**Table S5.** Number of matched walking bouts and strides in the real-life walking condition (RLW). Longitudinal within-subject comparison of bout length in real life (RLW) for baseline (BL) and 1-year follow-up assessment (FU) before and after pre-processing. p-values determined by Wilcoxon signed-rank test for both follow-up assessments relative to baseline. Shown are analyses for the whole group of degenerative cerebellar disease (DCD).

| RLW | Bout length (#Strides) | | p $\Delta$(FU-BL) | #Bouts | | #Strides | |
| --- | --- | --- | --- | --- | --- | --- | --- |
|  | BL | FU |  | BL | FU | BL | FU |
| Non-matched | 47.2±32.6 | 61.86±48.7 | 0.530 | 1450 | 1404 | 38507 | 53610 |
| Matched | 58.6±63.9 | 57.5±57 | **0.029** | 447 | 447 | 26872 | 26306 |

### Supplement E6 – Longitudinal within-subject comparison for the SCA1/2/3 subgroup

**Table S6.**
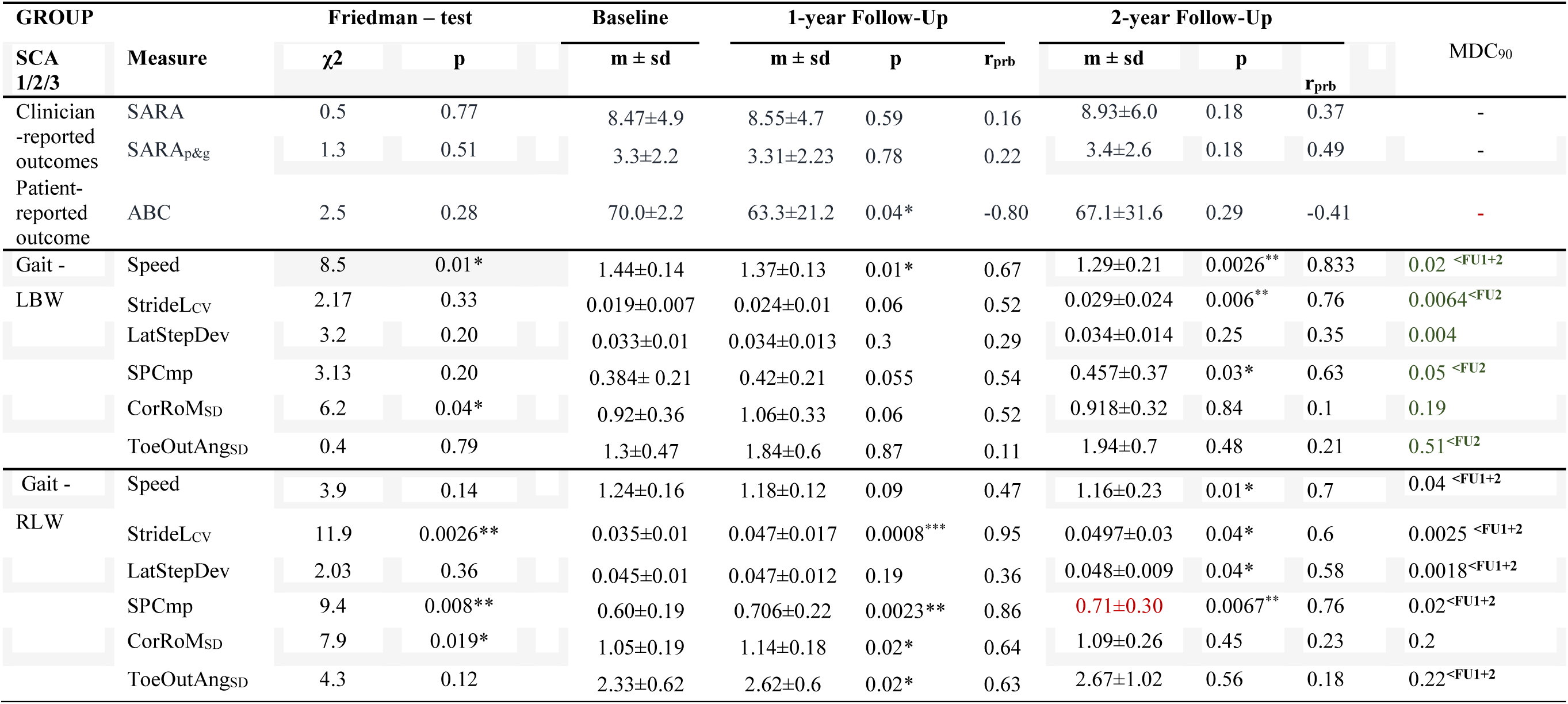
Longitudinal within-subject comparison for the SCA_1/2/3_ subgroup. Shown are results of clinical ataxia ratings (SARA score and SARA_p&g_ posture&gait subscore^41^) as well as of gait measures in clinical assessment (LBW) and in real life (RLW) for baseline, 1-year and 2-year follow-up assessments. Friedman test determined within-group longitudinal differences (^+^, p<0.1). Post-hoc test **p-**values determined by Wilcoxon signed-rank test for both follow-up assessments relative to baseline. Stars indicate significant differences between groups (*≡ p<0.05, **≡ p<0.0083 Bonferroni-corrected, ***≡ p<0.001). Effect sizes r_prb_ determined by matched-pairs rank-biserial correlation^51^. . MCD_90_ denotes the smallest reliable detectable change (90% confidence interval). The MDC column indicates whether this is smaller than the change between baseline and 1-year Follow-up (‘<FU1’) or/and between baseline and 2-year Follow-up (<FU1+2).

| GROUP |  | Friedman – test |  | Baseline | 1-year Follow-Up |  |  | 2-year Follow-Up |  |  | MDC<sub>90</sub> |
| --- | --- | --- | --- | --- | --- | --- | --- | --- | --- | --- | --- |
| SCA 1/2/3 | Measure | $\chi^2$ | p | m ± sd | m ± sd | p | $r_{prb}$ | m ± sd | p | $r_{prb}$ | |
| Clinician-reported outcomes | SARA | 0.5 | 0.77 | 8.47±4.9 | 8.55±4.7 | 0.59 | 0.16 | 8.93±6.0 | 0.18 | 0.37 | - |
|  | SARA<sub>p&g</sub> | 1.3 | 0.51 | 3.3±2.2 | 3.31±2.23 | 0.78 | 0.22 | 3.4±2.6 | 0.18 | 0.49 | - |
| Patient-reported outcome | ABC | 2.5 | 0.28 | 70.0±2.2 | 63.3±21.2 | 0.04* | -0.80 | 67.1±31.6 | 0.29 | -0.41 | - |
| Gait - | Speed | 8.5 | 0.01* | 1.44±0.14 | 1.37±0.13 | 0.01* | 0.67 | 1.29±0.21 | 0.0026** | 0.833 | 0.02 <FU1+2 |
| LBW | StrideL<sub>CV</sub> | 2.17 | 0.33 | 0.019±0.007 | 0.024±0.01 | 0.06 | 0.52 | 0.029±0.024 | 0.006** | 0.76 | 0.0064 <FU2 |
|  | LatStepDev | 3.2 | 0.20 | 0.033±0.01 | 0.034±0.013 | 0.3 | 0.29 | 0.034±0.014 | 0.25 | 0.35 | 0.004 |
|  | SPCmp | 3.13 | 0.20 | 0.384± 0.21 | 0.42±0.21 | 0.055 | 0.54 | 0.457±0.37 | 0.03* | 0.63 | 0.05 <FU2 |
|  | CorRoM<sub>SD</sub> | 6.2 | 0.04* | 0.92±0.36 | 1.06±0.33 | 0.06 | 0.52 | 0.918±0.32 | 0.84 | 0.1 | 0.19 |
|  | ToeOutAng<sub>SD</sub> | 0.4 | 0.79 | 1.3±0.47 | 1.84±0.6 | 0.87 | 0.11 | 1.94±0.7 | 0.48 | 0.21 | 0.51 <FU2 |
| Gait - | Speed | 3.9 | 0.14 | 1.24±0.16 | 1.18±0.12 | 0.09 | 0.47 | 1.16±0.23 | 0.01* | 0.7 | 0.04 <FU1+2 |
| RLW | StrideL<sub>CV</sub> | 11.9 | 0.0026** | 0.035±0.01 | 0.047±0.017 | 0.0008*** | 0.95 | 0.0497±0.03 | 0.04* | 0.6 | 0.0025 <FU1+2 |
|  | LatStepDev | 2.03 | 0.36 | 0.045±0.01 | 0.047±0.012 | 0.19 | 0.36 | 0.048±0.009 | 0.04* | 0.58 | 0.0018 <FU1+2 |
|  | SPCmp | 9.4 | 0.008** | 0.60±0.19 | 0.706±0.22 | 0.0023** | 0.86 | 0.71±0.30 | 0.0067** | 0.76 | 0.02 <FU1+2 |
|  | CorRoM<sub>SD</sub> | 7.9 | 0.019* | 1.05±0.19 | 1.14±0.18 | 0.02* | 0.64 | 1.09±0.26 | 0.45 | 0.23 | 0.2 |
|  | ToeOutAng<sub>SD</sub> | 4.3 | 0.12 | 2.33±0.62 | 2.62±0.6 | 0.02* | 0.63 | 2.67±1.02 | 0.56 | 0.18 | 0.22 <FU1+2 |

### Supplement E7 – Longitudinal within-subject comparison in the non-matched condition

**Table S7.** Longitudinal within-subject comparison for the group of degenerative cerebellar disease (DCD in the **non-matched condition**. Shown are gait measures in real life (RLW) for baseline, 1-year, and 2-year follow-up assessments. Friedman test determined within-group longitudinal differences (^+^, p<0.1). Post-hoc test **p-**values determined by Wilcoxon signed-rank test for both follow-up assessments relative to baseline. Stars indicate significant differences between groups (*≡ p<0.05, **≡ p<0.0083 Bonferroni-corrected, ***≡ p<0.001). Effect sizes r_prb_ determined by matched-pairs rank-biserial correlation^51^.

| non-matched |  | Friedman |  | Baseline | 1-year Follow-Up |  |  | 2-year Follow-Up |  |  |
| --- | --- | --- | --- | --- | --- | --- | --- | --- | --- | --- |
| | Measure | $\chi^2$ | p | m ± sd | m ± sd | p | $r_{prb}$ | m ± sd | p | $r_{prb}$ |
| **Gait -** | Speed | 2.1 | 0.33 | 1.22±0.14 | 1.19±0.13 | 0.33 | 0.23 | 1.17±0.21 | 0.02* | 0.54 |
| **RLW** | StrideLcv | 8.4 | 0.01<sup>+</sup> | 0.036±0.013 | 0.04±0.015 | 0.01* | 0.59 | 0.047±0.032 | 0.01* | 0.58 |
| | LatStepDev | 10.8 | 0.006<sup>+</sup> | 0.044±0.01 | 0.046±0.010 | 0.07 | 0.42 | 0.047±0.009 | 0.10 | 0.39 |
| | SPCmp | 14.9 | 0.0005<sup>+</sup> | 0.57±0.16 | 0.63±0.2 | 0.0089** | 0.62 | 0.71±0.40 | 0.01* | 0.58 |
| | CorRoM<sub>SD</sub> | 6.8 | 0.03<sup>+</sup> | 1.01±0.19 | 1.08±0.19 | 0.01* | 0.60 | 1.05±0.21 | 0.32 | 0.24 |
| | ToeOutAngle<sub>SD</sub> | 8.2 | 0.01<sup>+</sup> | 2.35±0.6 | 2.57±0.58 | 0.0081** | 0.63 | 2.61±0.73 | 0.03* | 0.52 |

### Supplement E8 – Correlations of gait measures between lab-based walking and real-life walking

**Table S8.** Correlations between gait measures in conditions LBW and RLW are given for the DCD group. Effect sizes of correlations are given using Spearman’s ρ. (*≡ p<0.05, **≡ p<0.0083 Bonferroni-corrected, ***≡ p<0.001).

| Measure | Corr LBW, RLW |  |
| --- | --- | --- |
| | $\rho$ | p |
| Speed | 0.7 | 0.006** |
| StrideLcv | 0.81 | <0.001*** |
| LatStepDev | 0.74 | 0.002** |
| SPCmp | 0.74 | 0.002** |
| CorRoM<sub>SD</sub> | 0.74 | 0.002** |
| ToeOutAng<sub>SD</sub> | 0.23 | 0.40 |

